# Automated aortoiliac calcification segmentation from arterial-phase computed tomography scans: development and validation

**DOI:** 10.64898/2026.11.19.26368507

**Authors:** Benjamin C. Liu, Andrea T. Fisher, Jillian Melbourne, Arash Fereydooni, Pauline Berens, Anoushka Lakshmi, Malte E.K. Jensen, Fatima Rodriguez, Akshay S. Chaudhari, Shipra Arya

## Abstract

**Introduction:** Arterial calcium is widely used as a biomarker for coronary artery disease but is underutilized in peripheral artery disease (PAD) due to challenges in automated measurement of peripheral vascular calcium. We developed a fully automated deep-learning pipeline for measurement of aortoiliac calcium from arterial-phase, contrast-enhanced abdominopelvic computed tomography angiography (CTA) to extract this important PAD biomarker.

**Methods:** The pipeline uses two nnU-Net models for aortoiliac arterial segmentation and identification of calcified voxels. We trained on 56 computed tomography (CT) scans and validated on 106 scans by comparing the pipeline’s calcium estimation from CTA against calcium burden on corresponding non-contrast phase CT within manually segmented arterial contours across a range of radiodensity thresholds (100–1000 Hounsfield units [HU]). Using the non-contrast HU threshold that maximized correlation between measures, we performed linear regression with five-fold cross-validation between pipeline-estimated and non-contrast calcium quantities, and risk reclassification analysis across quartiles using Monte Carlo simulations to compare pipeline and non-contrast-derived calcium burden quartiles while measuring quadratically weighted kappa.

**Results:** Non-contrast calcium burden calculated from voxels with attenuation ≥400 HU correlated best with CTA calcium burden (Pearson r=0.952 for aortic calcium; Pearson r=0.955 for iliofemoral calcium) and was used for subsequent analysis. Five-fold cross-validation of non-contrast calcium volume prediction from pipeline-estimated CTA calcium burden, using ≥400 HU to define non-contrast calcium voxels, demonstrated aggregated Pearson r>0.91 for all held-out folds in all three arterial regions tested. In risk reclassification analysis, quadratically weighted kappa for agreement between predicted and ground truth calcium quartile classifications was κ=0.88 for the aorta, κ=0.84 for bilateral iliofemoral arteries, and κ=0.80 for all regions.

**Conclusion:** Aortoiliac calcium burden can be accurately quantified from CTA with excellent reproducibility against calcium voxels ≥400 HU on non-contrast imaging. Quantification of this important biomarker expands opportunities for improved peripheral vascular disease diagnosis and risk prediction.

**Graphical Abstract:** 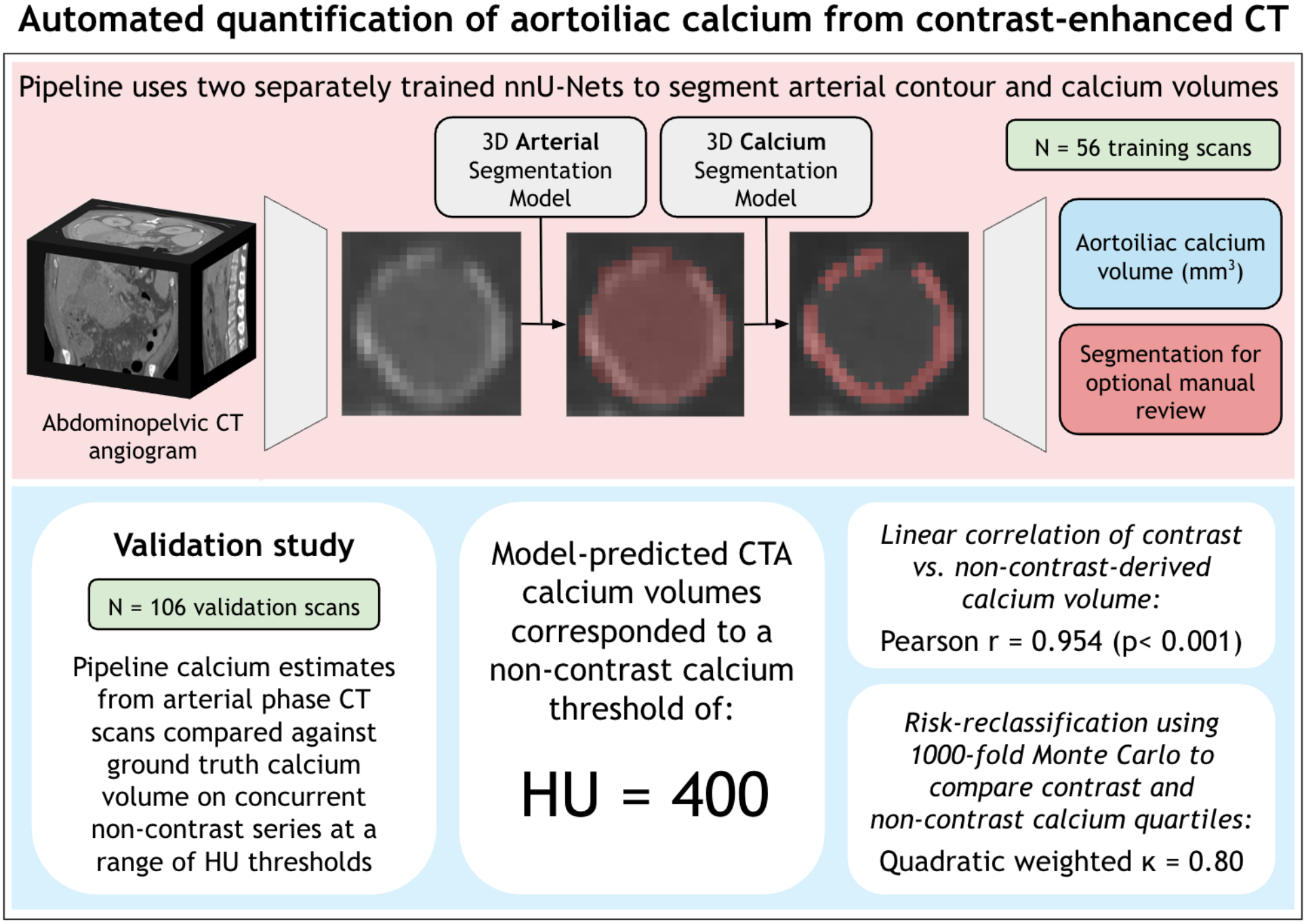

## Introduction

Peripheral artery disease (PAD) affects approximately 200 million people worldwide.[1] Although mild to moderate PAD usually presents as claudication, advanced manifestations of the disease can include chronic wounds and tissue loss necessitating amputation.[2] Vascular providers have treatment strategies for all clinical severity levels of PAD,[3-5] but patients are often not referred for vascular evaluation until their disease is advanced.[6-8]

Coronary artery disease (CAD) shares pathophysiology and radiographic features with PAD,[9] and the computed tomography (CT)-derived coronary artery calcium (CAC) score has been successfully developed to facilitate early CAD diagnosis and risk stratification.[10-13] We therefore aim to create a similar CT-derived, fully automated peripheral artery calcium score (PACS) to aid in PAD diagnosis and risk stratification,[14] starting with quantification of aortoiliac calcium burden. Calcium deposits in the aortoiliac vessels are a hallmark of aortoiliac occlusive disease[15] and contribute to a reduced ankle-brachial index (ABI), which is a key diagnostic measure of PAD.[16] In patients with PAD, aortoiliac calcium burden is associated with major adverse limb events (MALE) at all time points[17, 18] and particularly after aortoiliac revascularization.[19, 20] Even for those without PAD, aortoiliac calcification predicts major adverse cardiovascular events (MACE).[21, 22]

To facilitate integration of PACS into existing PAD diagnosis and treatment workflows, we extracted calcium from arterial-phase, contrast-enhanced CT angiography (CTA) scans since this protocol is used by vascular providers to evaluate PAD from the aortoiliac segment through the tibial vessels.[23] Past work quantifying aortoiliac calcium was typically performed on non-contrast-enhanced CT using a modified version of the coronary calcium Agatston score,[24-27] although manual techniques exist to extract an aortic calcium score from CTA scans as well.[28] Two previous studies detail development of neural networks for automatic aortic calcium extraction from CTA scans, but neither is validated against non-contrast ground truth; instead, both are validated against clinician hand-drawn segmentations of calcium volume on the same CTA scans.[29, 30] Because clinician-generated calcium segmentations from CTA scans can be subjective, the external validity of these studies is limited. We propose validation against calcium volume extracted from non-contrast CT because grounding against an objective metric makes our technique and resulting clinical risk metrics replicable for patients at other institutions.

Here, we introduce a fully automated pipeline for quantifying aortoiliac calcium burden in arterial-phase, contrast-enhanced CT scans that is validated against threshold-defined non-contrast calcium volumes.

## Methods

### Deep Learning System

We designed an automated deep learning framework to segment and quantify aortoiliac calcium from CTA scans. Our pipeline integrates two fully supervised three-dimensional (3D) nnU-Net segmentation models: the arterial segmentation model (ART nnU-Net) segments the aortoiliac vasculature, and the calcium segmentation model (CA nnU-Net) segments calcium within the arterial region. Calcium segmentations are then volumetrically quantified (Figure 1). The techniques used to generate training masks are not used in the final, fully automated pipeline, which consists only of nnU-Net segmentation. Training masks for both models were generated slice-by-slice in 2D and subsequently stacked into 3D volumes, which were then used to train two fully 3D nnU-Net models (ART and CA nnU-Net) that are applied together during inference.

**Figure 1.**
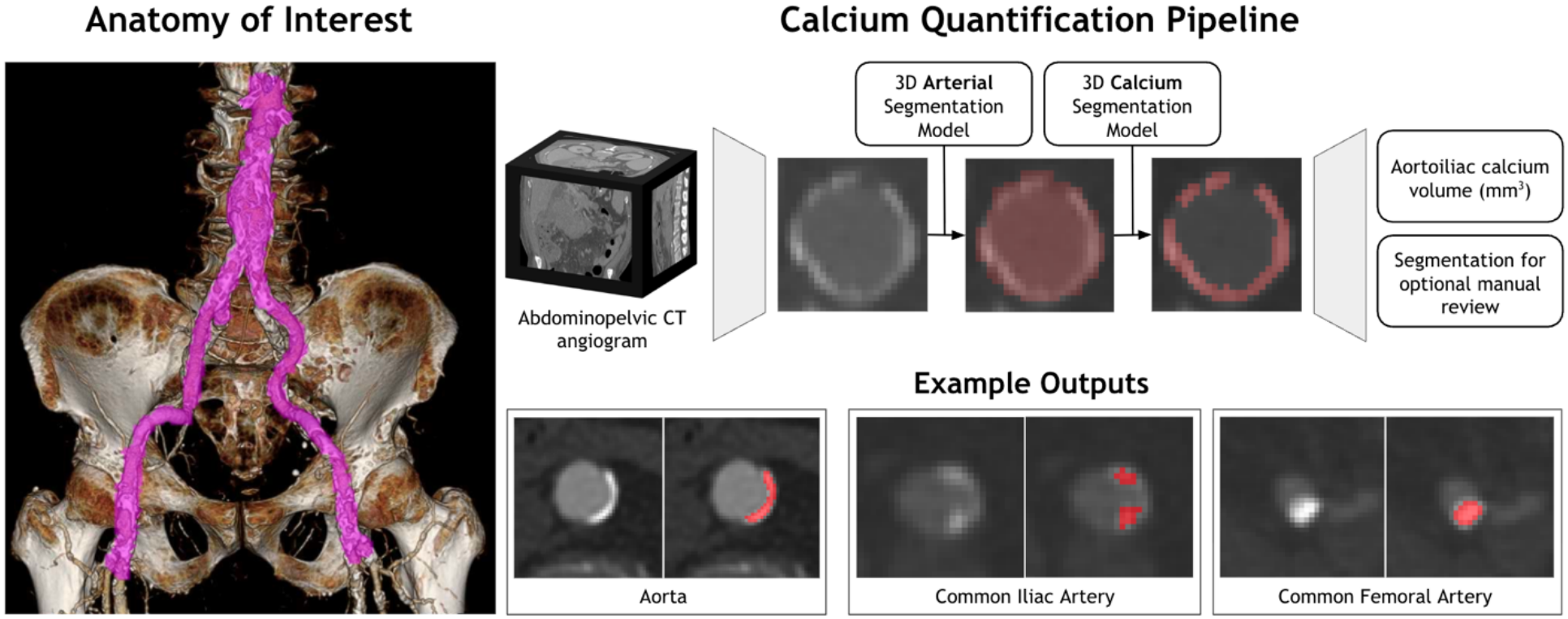
(Left) Anatomic region of calcium quantification highlighted, including the abdominal aorta, common and external iliac arteries, and common femoral arteries terminating at the common femoral artery bifurcation into the profunda femoris and superficial femoral arteries. (Right) Pipeline overview, which sequentially uses two fully supervised three-dimensional segmentation models to predict aortoiliac calcium volume from an abdominopelvic CT angiogram, and example calcium segmentations.

### Dataset Acquisition and Allocation

To create and evaluate our pipeline, we obtained CTA scans that included the abdomen, pelvis, and bilateral lower extremities (CTA runoff). Our institutional protocol for these studies includes a non-contrast phase of the abdomen and pelvis. Using Current Procedural Terminology (CPT) code 75635, we identified all patients in our hospital’s electronic health record research database who had undergone CTA runoff between 2006 and 2024. Separate, non-overlapping subsets of these data were compiled for training and validation. Institutional review board (IRB) approval for this retrospective study was obtained from our institution (IRB reference number 66789). The requirement for informed consent was waived by the institutional review board. Validation used paired arterial-phase and non-contrast series from the same examination.

ART and CA nnU-Net training scans were curated to ensure generation of high-quality training masks using our adapted Segment Anything Model 2 (SAM-2)[31] and calcium maximum likelihood estimation techniques (Supplementary Section). Both training sets were drawn from a pool of scans unlikely to have arterial occlusions using a text-matching approach against radiology reports. For ART nnU-Net training, we curated a set of 52 scans with no intraarterial devices, surgical bypasses, anatomic abnormalities, or pseudoaneurysms with contiguous, well-opacified arteries. For CA nnU-Net, we selected a set of 50 scans that had no intraarterial devices, surgical bypasses, anatomic abnormalities, or pseudoaneurysms, and we sought a diversity of calcification quantities and calcification anatomic distributions (Figure 2). After generation of training segmentations, 14 and 16 scans were eliminated from ART and CA nnU-Net training pools, respectively, due to insufficient accuracy of segmentations on visual inspection. The final training cohorts were 38 scans for ART nnU-Net and 34 scans for CA nnU-Net, of which 16 scans were shared between cohorts. Training cohort sizes were chosen by evaluating trained nnU-Net performance with increasing amounts of training data until performance was satisfactory.

**Figure 2.**
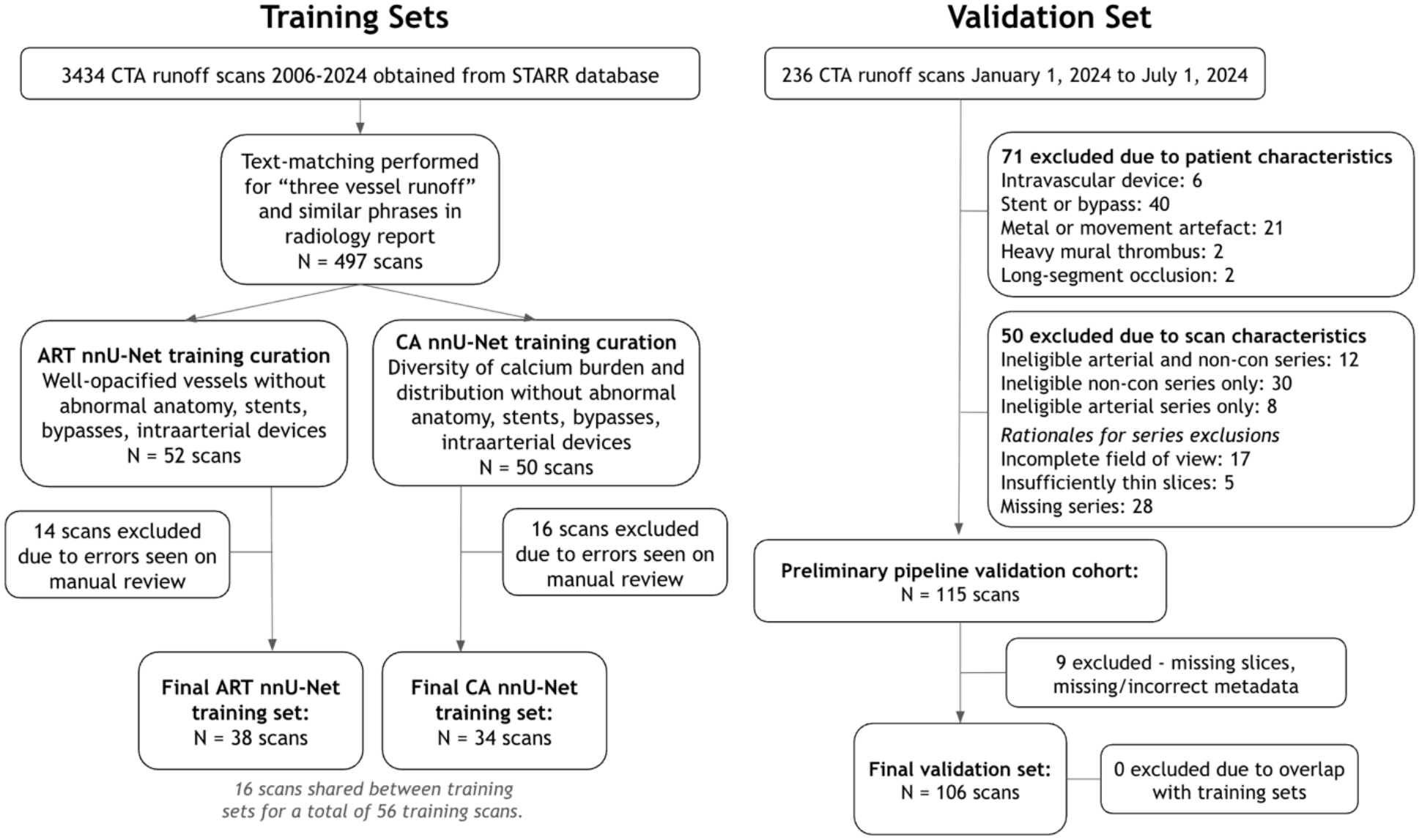
Flow diagrams for creation of training and validation datasets. CTA runoff = Arterial-phase computed tomography angiography of the abdomen through the lower extremities ART nnU-Net = Arterial segmentation nnU-Net model CA nnU-Net = Calcium segmentation nnU-Net model

To validate contrast-enhanced calcium scores from the full pipeline, we reviewed all CTA runoff scans acquired from January 1, 2024, to July 1, 2024, at our institution and added scans meeting inclusion criteria to the validation set. This collection period was chosen to ensure use of modern scanners with modern imaging protocols. Scans were excluded if they contained femoral intravascular devices, stents, or arterial bypasses; showed motion or metal artifacts; did not cover the region from the L1 superior endplate through the common femoral artery bifurcations; or did not include both non-contrast and arterial phases reconstructed at a slice thickness ≤2.5 mm (Figure 2). Intermediate vessel segmentations of aortic aneurysms with significant mural thrombus extended only to the bounds of the arterial blood pool and not to the arterial wall (Supplemental Figure S1A). These were therefore excluded (2 scans). Short-segment (< 3 cm) arterial occlusions were included; manual review of these scans revealed successful calcium detection even at the margin of fully thrombosed vessels (Supplemental Figure S1B). Long-segment arterial occlusions were excluded. Several scans had corrupted image data (missing axial slices in the Digital Imaging and Communications in Medicine [DICOM] series, incorrect/missing CT metadata); scans not passing completeness/quality-control checks were also excluded. None of the training scans were included in the validation set (N=106; Figure 2).

Training and validation scan characteristics and basic demographics are available in Supplemental Table S1.

### Arterial Segmentation Training Masks

To segment the aortoiliac vasculature, we trained ART nnU-Net using algorithmically generated segmentation masks subject to manual verification. We defined the target vessels as the aorta from the superior endplate of L1 to the bilateral common femoral artery bifurcations (Figure 1). To generate training masks, we leveraged the open-source SAM-2 from Meta AI,[31] which is a general-purpose segmentation tool that maintains object consistency across frames by integrating real-time predictions with a memory of prior states. TotalSegmentator[32] was used to initialize an aortic mask at the L1 superior endplate, and segmentation was propagated down the arterial tree using SAM-2 (v2.1 Hiera-Large checkpoint; Figure 3). See the Supplementary Section for further detail.

**Figure 3.**
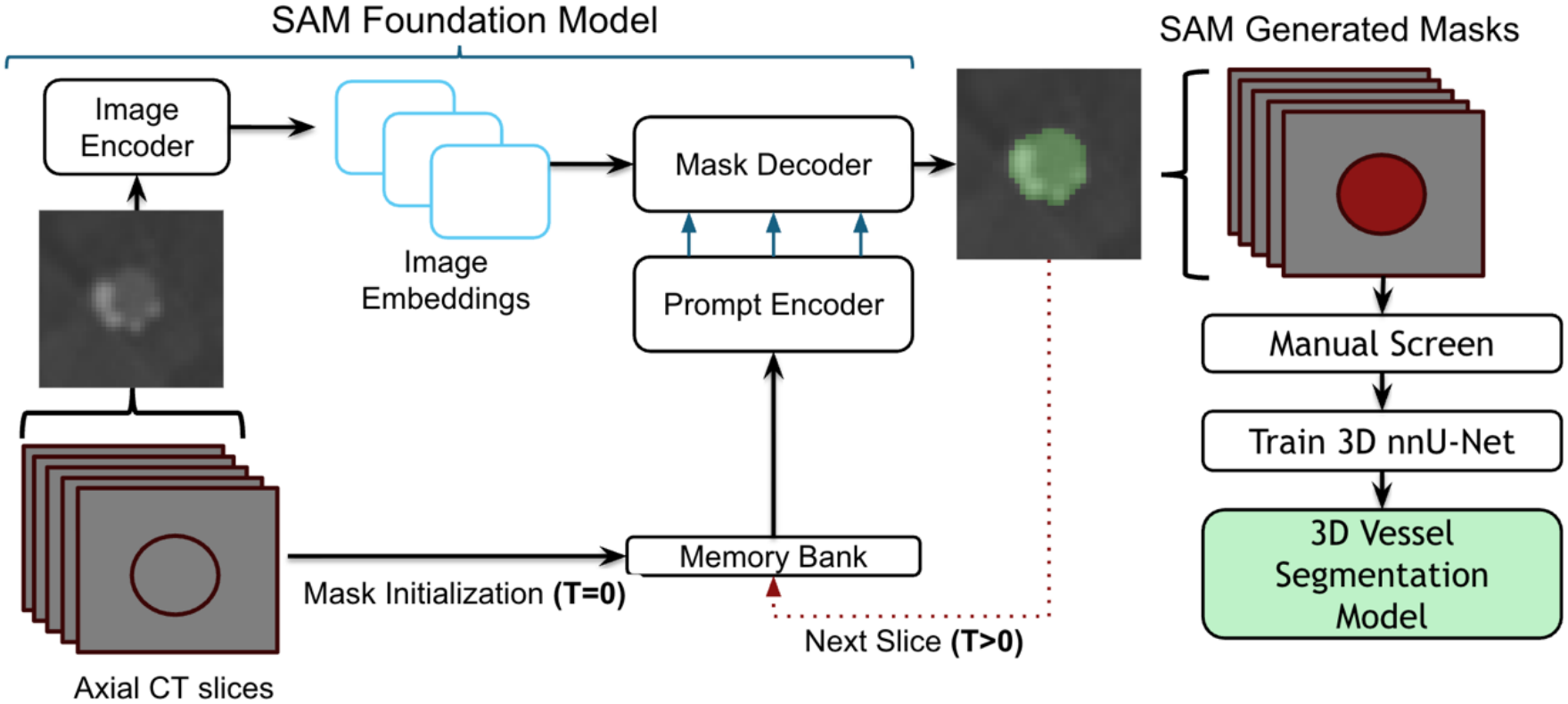
Mask generation process for training ART nnU-Net. 3D vessel training masks were generated by adapting the SAM-2 video segmentation method. Acceptable masks following manual screening were used to train ART nnU-Net.

Following training mask generation, a vascular surgeon (A.T.F.) manually reviewed each mask and eliminated any masks in which the segmentation did not match the visible bounds of the arteries. The final labeled dataset was used to train ART nnU-Net.

### Calcium Segmentation Training Masks

To generate training masks for CA nnU-Net, we developed an automated labeling pipeline grounded in the differential radiodensity of calcium relative to patient-specific anatomic landmarks (Figure 4). Our method leveraged the observation that aortoiliac calcium intensity closely resembles that of the dense lower spine, distinguishing it from the baseline intensity of the luminal blood pool even in the presence of arterial contrast. TotalSegmentator (v2)[32] was used to identify the T12–L5 vertebrae and the aorta, and the radiographic intensity distributions of bone and the central aortic lumen were modeled. ART nnU-Net was used to generate arterial segmentations and define the set of calcium candidate voxels for each scan. Candidate voxels within the arterial segmentation were then compared against the bone and blood pool distributions to classify bone-similar voxels as calcium, and watershed mask dilation was applied to yield final arterial calcium masks. See the “Generation of Training Data” subsections in the Supplementary Section for further detail.

**Figure 4.**
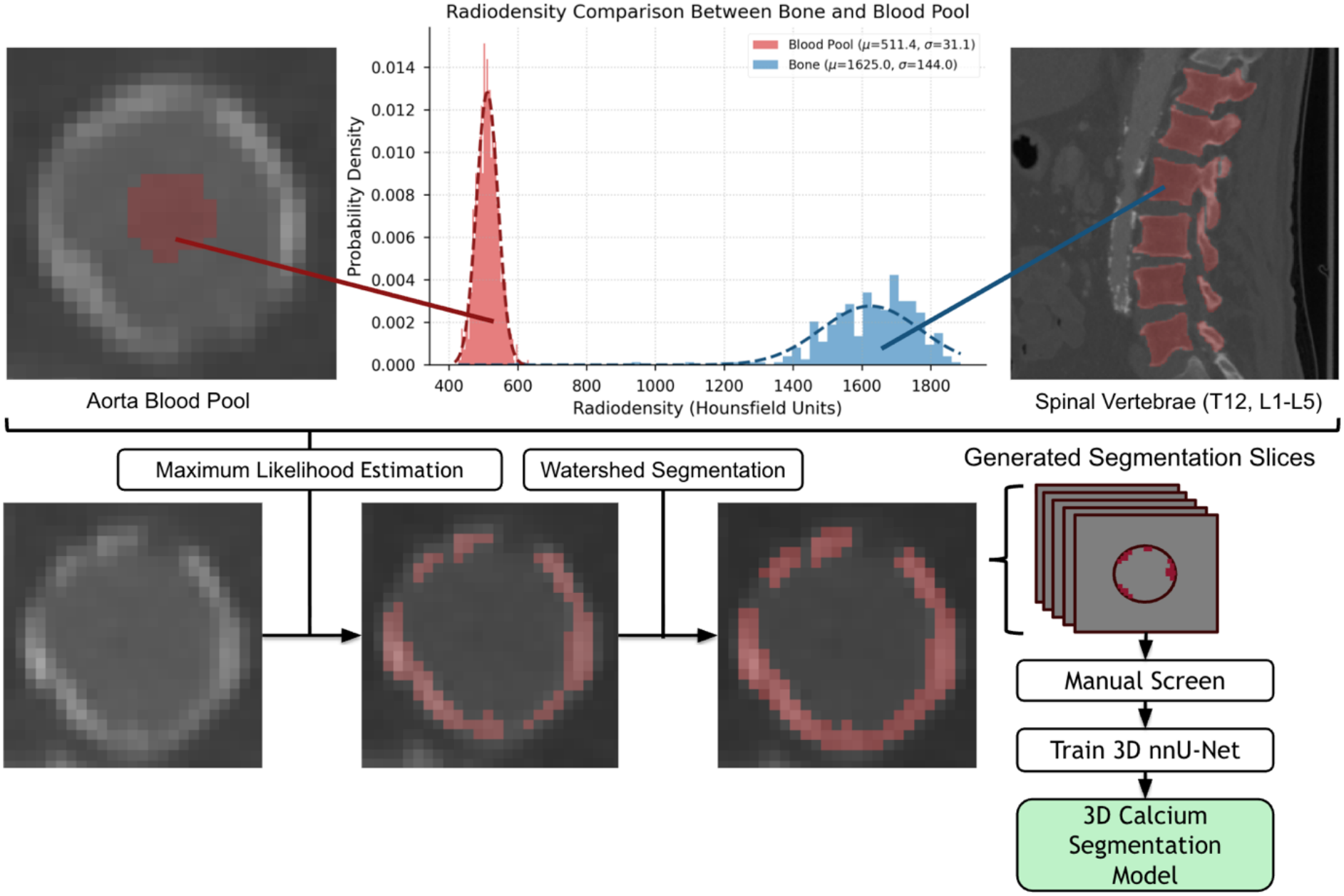
Mask generation process for training CA nnU-Net. Initial calcium region-of-interest (ROI) predictions were generated with a maximum likelihood estimation method that assesses pixels relative to radiodensity distributions of the spinal vertebrae versus the aortic blood pool (top). Watershed segmentation was conducted to expand the initial predictions to neighboring calcium voxels. Acceptable segmentations following manual screening were used to train CA nnU-Net.

After automated calcium labeling of each arterial-phase series, a vascular surgeon (A.T.F.) manually reviewed each calcium mask to eliminate masks in which any segmentation did not correspond with visible calcium. The final labeled dataset was used to train CA nnU-Net.

### Arterial and Calcium Segmentation Model Training

We trained both ART nnU-Net and CA nnU-Net (nnU-Net v2) with an 80:20 training/validation split applied to both the arterial and calcium nnU-Net training cohorts (31:7 and 28:6). We utilized a stochastic gradient descent (SGD) optimizer with Nesterov momentum (μ=0.99) and weight decay (3 × 10^−5^), paired with a polynomial learning rate scheduler (initial learning rate η=0.01). Training proceeded for 1,000 epochs with a batch size of 2 and a patch size of 96×96×256 voxels. We optimized an equally weighted composite objective function summing soft Dice and cross-entropy loss and selected our best model based on the highest validation Dice score. All experiments were executed on a single NVIDIA RTX A4000 graphics processing unit (GPU), with each training run converging within 36 hours.

### Validation using non-contrast calcium ground truth

To evaluate pipeline performance on CTA scans, we used each corresponding non-contrast phase from the same imaging study to generate ground truth calcium volume scores. For each non-contrast abdominopelvic CT phase, a vascular surgery resident (A.T.F.) manually segmented the contours of the abdominal aorta, bilateral common iliac arteries, bilateral external iliac arteries, and bilateral common femoral arteries. The proximal extent of segmentation was defined by the superior endplate of the L1 vertebra, and the distal extent was defined by the common femoral artery bifurcation on each side. The aorta, right iliac system, and left iliac system were segmented separately to facilitate comparisons by anatomic region. For the purposes of this study, each common femoral artery was considered part of the ipsilateral iliac system. A filter was applied to each annotated arterial region of the non-contrast CTs to quantify voxels at each intensity in Hounsfield units (HU). This was used to generate curves depicting a distribution of voxels with intensity at or above each threshold, returning differing amounts of calcium at different minimum intensity thresholds. Thresholds were increased from HU = 100 to HU = 1000 at a granularity of 20 HU.

### Statistical Analysis

All statistical analyses were performed using Python 3.8 with NumPy 1.18, SciPy 1.5, Pandas 1.1, scikit-learn 0.23, and Matplotlib 3.2. Non-contrast calcium volumes at each threshold were plotted against pipeline-extracted CTA calcium volume to calculate the Pearson correlation coefficient at each threshold, and the threshold that maximized the Pearson correlation coefficient was selected for subsequent analysis.

To visualize generalization of derived linear regression equations to unseen samples, we performed five-fold cross-validation for each arterial configuration (aortic, bilateral iliac, all three regions). For each fold, the held-out set was evaluated on the linear regression conversion derived from the other four sets to assess how closely the non-contrast volume predicted from CTA volume aligned with ground truth non-contrast calcium volume at the selected threshold.

Risk reclassification analysis was then performed using 1,000 Monte Carlo simulations, in each of which 80% of the data were randomly selected to derive the linear conversion parameters and the remaining 20% were evaluated as a held-out set. We determined whether patients classified in each risk stratum based on non-contrast-derived calcium volume remained in the same risk stratum using the linear conversion applied to CTA-derived calcium volume. Unlike the coronary artery Agatston score,[33] there is no accepted risk stratification schema for aortoiliac calcium. We therefore defined risk strata by dividing both the non-contrast and CTA calcium volume scores into quartiles.

## Results

### Study Population

To train both ART nnU-Net and CA nnU-Net, we curated 38 (29,922 slices) and 34 scans (28,118 slices), respectively. Since 16 patients were shared between ART nnU-Net and CA nnU-Net training sets, there were 56 training scans. We then validated the integrated aortoiliac calcium segmentation pipeline on a non-curated set of 106 sequential CT scans (169,942 slices) meeting inclusion criteria. In the ART nnU-Net, CA nnU-Net, and validation cohorts, female patients accounted for 14/38 (37%), 11/34 (32%), and 67/106 (63%), respectively. Mean ages (standard deviations) were 65.5 (14.3), 68.8 (9.6), and 71.2 (17.6) years in the ART, CA, and validation cohorts, respectively. Scan characteristics are detailed in Table S1.

### Arterial and Calcium Segmentation Model Results

The deployed ART nnU-Net and CA nnU-Net models achieved validation Dice scores of 0.929 and 0.826, respectively, and exhibited stable convergence without evidence of overfitting.

### Agreement Between Methods

Following model development, we evaluated our integrated calcium segmentation pipeline on the validation cohort of 106 patients. We compared pipeline-quantified calcium from arterial phase with corresponding non-contrast phase calcium volumes across a variety of thresholds. HU thresholds were applied uniformly to the vessels of interest in a non-contrast phase to derive ground truth calcium volume measurements. Defining non-contrast calcium as voxels with attenuation ≥400 HU within the arterial contour corresponded best with pipeline-quantified calcium from arterial phase (Figure 5).

**Figure 5.**
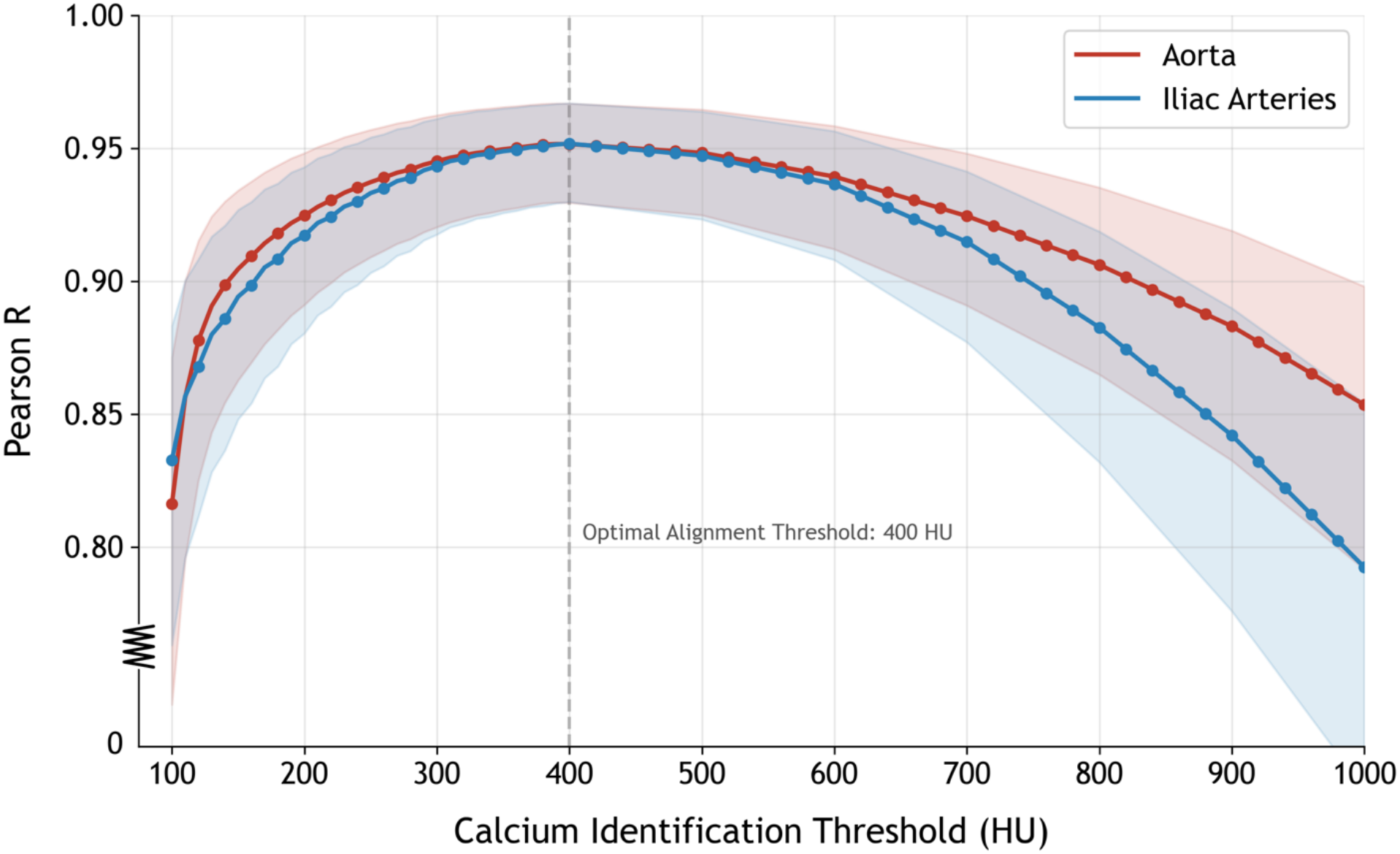
Distribution of Pearson correlation coefficients (r) between predicted calcium volume (mm^3^) and ground truth calcium volume (mm^3^) across different calcium identification thresholds (HU) used for ground truth classification. Comparisons were stratified by the aorta (red) and iliac arteries (blue); shaded areas indicate 95% confidence intervals.

Linear correlations between non-contrast phase-derived calcium volume using a calcium voxel definition of ≥400 HU and arterial phase-extracted calcium volume were statistically significant for all anatomic configurations (aortic Pearson r=0.952, R^2^=0.905, p<0.001; bilateral iliac Pearson r=0.955, R^2^=0.912, p<0.001; all three regions Pearson r=0.954, R^2^=0.910, p<0.001). Setting non-contrast phase-derived calcium volume as y and arterial phase-extracted calcium volume as x, the linear equations were y=1.19x+334.76 for aorta, y=1.25x+403.16 for bilateral iliac, and y=1.22x+711.70 for all regions. Bland-Altman curves demonstrated that CTA predictions tended to underestimate calcium in the iliac arteries for larger calcium burdens (Figure 6).

**Figure 6.**
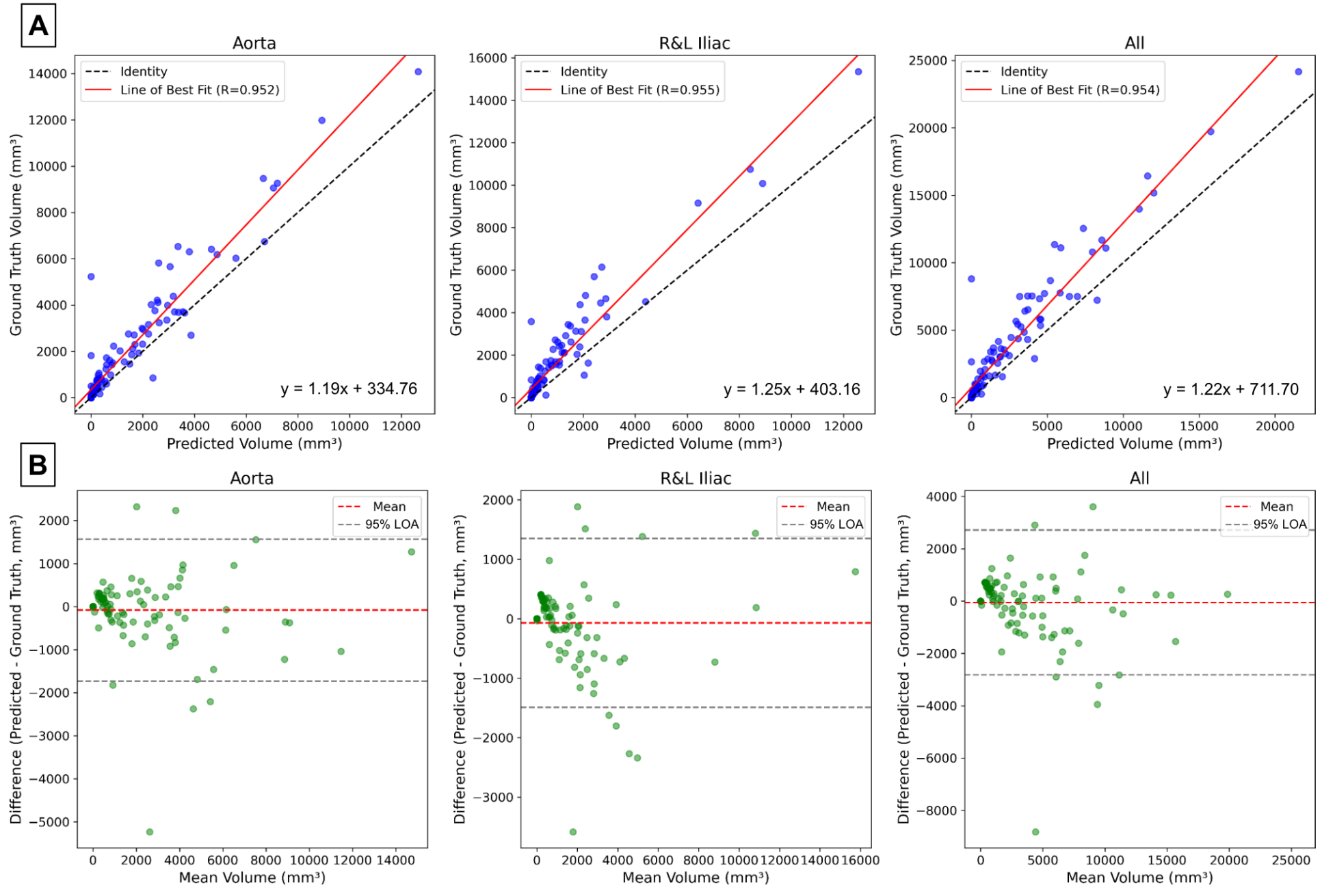
(A) Linear plots between arterial phase-extracted and non-contrast phase-derived calcium volume using a non-contrast calcium voxel definition of ≥400 HU for each anatomic region. Correlation analyses were performed for aorta, bilateral iliac arteries, and all three regions. Linear equations corresponding to the red line are at the bottom right of each plot. (B) Bland-Altman plots for each anatomic region. The dashed red line indicates the mean difference between prediction and ground truth; the dashed gray lines indicate 95% limits of agreement.

Five-fold cross-validation was performed to predict non-contrast calcium volume from arterial-phase-extracted calcium volume, with non-contrast calcium voxels defined as those with attenuation ≥400 HU. Derived linear relationships with held-out set predictions from the best-performing fold for each arterial configuration are demonstrated in Figure 7. Across all folds and arterial configurations, we observed strong agreement between model predictions and ground truth with aggregated Pearson r>0.91 and R^2^>0.84 (p<0.01) in a 5-fold cross-validation assessment.

**Figure 7.**
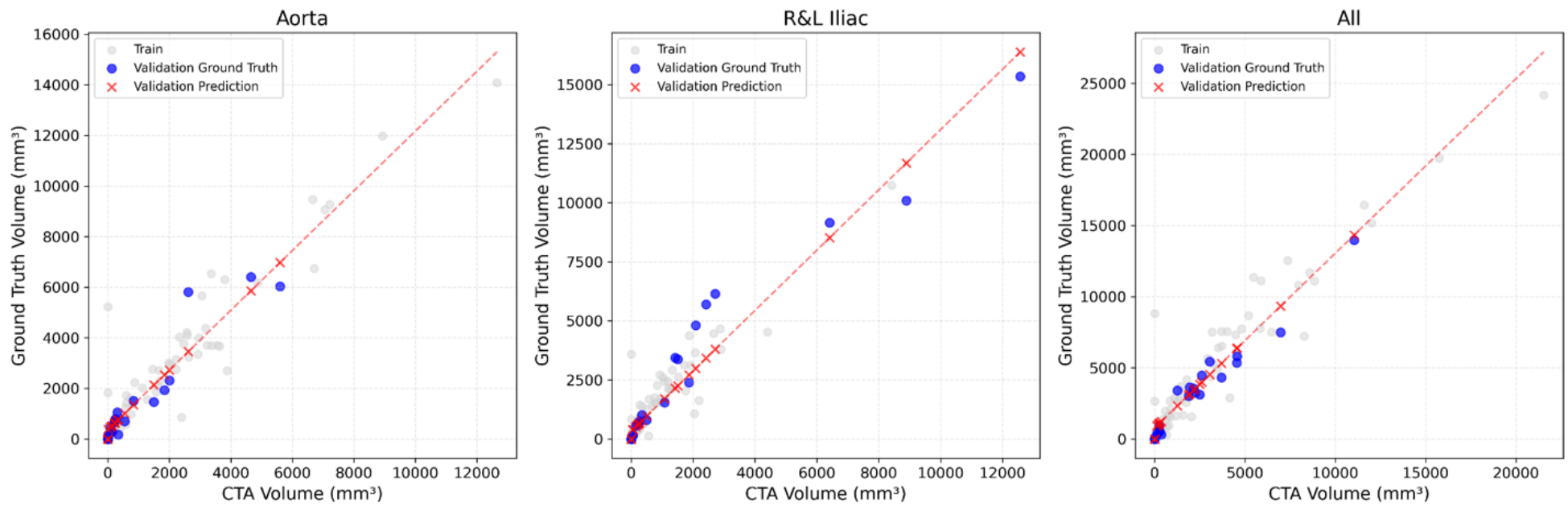
Visualizations of differences between CTA-derived calcium score predictions (red X symbols, linear regression-derived) and corresponding ground truth scores (blue dots, vertically adjacent).

We then performed risk reclassification analysis to determine whether patients classified into a given risk category using non-contrast phase-derived calcium volume remained in that same risk category using the linear conversion applied to CTA-derived calcium volume. We divided both arterial and non-contrast calcium volumes into quartiles to define risk categories, in the absence of accepted aortoiliac or abdominal aortic calcium risk classification schemata. When the raw CTA pipeline calcium volume was zero, we set the predicted non-contrast calcium volume to zero rather than adding the regression intercept, thereby avoiding systematic overprediction for scans without visible calcium. Quadratically weighted kappa was calculated for each anatomic configuration, with aortic κ=0.88 (95% CI 0.68–0.98), bilateral iliac κ=0.84 (95% CI 0.61–0.95), and all regions κ=0.80 (95% CI 0.57–0.94). Results for the 20% validation split for each Monte Carlo fold were accumulated into heatmaps (Figure 8). Non-contrast calcium volume cutoffs at the 25th, 50th, and 75th percentiles were 148/855/3118 mm^3^ (aorta), 36/642/2130 mm^3^ (bilateral iliac), and 296/1815/5607 mm^3^ (combined).

**Figure 8.**
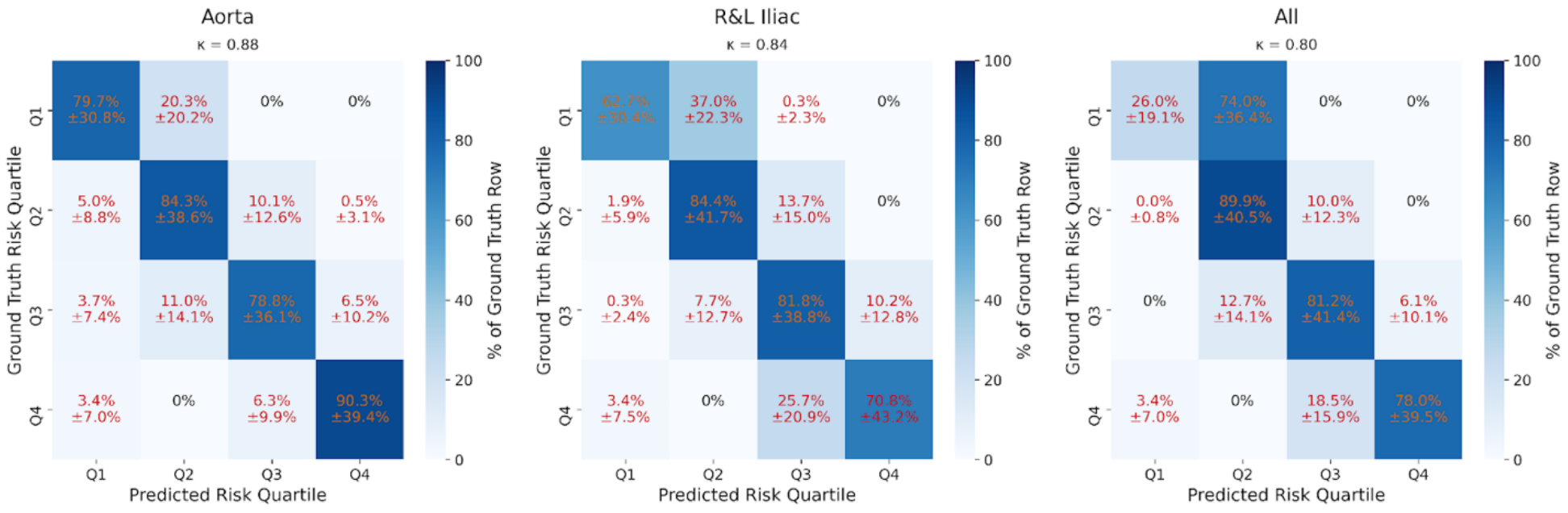
Agreement between CTA-predicted (columns) and non-contrast (rows) calcium burden quartiles for aorta, bilateral iliac arteries, and all regions (non-contrast threshold ≥400 HU). Row percentages: mean ± SD across 1,000 random 80:20 training/validation splits. Quartile thresholds were derived from each training set. κ denotes quadratically weighted kappa.

## Discussion

In this study, we developed and validated a fully automated pipeline for quantification of aortoiliac calcium volume from CTA scans. Calcium burden extracted from CTA scans with this technique reliably corresponds with voxels ≥400 HU on non-contrast scans, with Pearson r of >0.95 for aortic, iliac, and aortoiliac segments. Derived linear equations can be used to relate CTA-extracted calcium to non-contrast phase-derived calcium using a calcium voxel definition of ≥400 HU. Risk reclassification analysis demonstrated strong agreement between CTA-derived calcium volume quartile and non-contrast ground truth calcium volume quartile, with κ≥0.80 for aortic, iliac, and aortoiliac segments. Validation analyses were based on similar previous research on coronary artery calcium.[34, 35] Our technique for automatic CTA-derived aortoiliac calcium quantification is thus highly reliable compared to manual techniques applied to non-contrast CT scans.

Accurate, consistent quantification of aortoiliac calcium has multiple potential clinical applications. Since aortoiliac calcium is associated with PAD and reduced ABIs,[15, 16] opportunistic screening of abdominopelvic CTs could help identify patients with undiagnosed PAD[2, 17, 18] especially in instances where traditional ABI measurement is artifactually elevated in calcified vessels.[36-40] The share of PAD patients with comorbidities that lead to calcified tibial vessels (diabetes, end-stage renal disease, advanced age)[36, 41, 42] is increasing,[43, 44] and a more robust diagnostic tool is necessary. Because heavy aortoiliac calcification also indicates elevated cardiovascular risk in general,[21, 22] early medical management and lifestyle changes could prevent progression to both major adverse limb and cardiovascular events in these patients. Recent society guidelines on the management of dyslipidemia recognize the growing importance of coronary calcifications as a prognostic factor;[13] exploration of aortic calcification may contribute further to prediction of long-term cardiovascular outcomes. Aortoiliac calcium quantification could also be used for preoperative risk stratification as abdominal surgeries are often planned with abdominal CT, from which an aortoiliac calcium score could be derived to identify patients who may benefit from further preoperative cardiac workup.

To our knowledge, this is the first study to validate fully automated CTA-derived aortoiliac calcium volumes against threshold-defined calcium volumes on paired non-contrast CT. Halkoaho et al. and Luo et al. previously used neural networks to segment aortoiliac calcium from arterial-phase CT, with manual plaque segmentations from the same contrast-enhanced images as reference standards.[29, 30] Our non-contrast reference uses HU thresholding within manually delineated arterial contours, without manual plaque segmentation. This anchors CTA validation to an attenuation-defined reference, reducing potential annotator error/bias in generating ground truth comparisons. We empirically identified ≥400 HU as the non-contrast calcium definition that best corresponded to CTA-derived volume, assessed volume agreement with Bland-Altman analysis, and evaluated volume conversion and calcium-burden quartile agreement using held-out data. Coverage through the common femoral artery bifurcations further supports assessment of the aortoiliofemoral territory relevant to PAD.

Although PACS extraction can be done with a simpler, threshold-based approach from non-contrast CT,[25, 45] our previous work demonstrated the limitations of a non-contrast approach. We used non-contrast-derived PACS to predict lower extremity amputation risk[26] but had to specifically request each non-contrast phase series to be extended through the bilateral lower extremities (as opposed to only in the abdominopelvic region) since this protocol is non-standard for a typical vascular CTA runoff study.[46] Requiring changes to typical radiology protocols would likely inhibit widespread adoption of PACS and raise concern for added radiation exposure. Automatic calcium extraction from CTA will instead allow for pipeline integration into existing vascular provider workflows. The next step will involve extending our method down the legs into the femoropopliteal and tibial regions. Full lower extremity calcium scoring may enable us to create a better diagnostic/prognostic tool for patients in whom accurate ABIs are not possible, and to better predict which intervention strategies are most likely to succeed for specific patients.

The threshold we derived for non-contrast calcium equivalence to CTA calcium exceeds the threshold used for Agatston scoring (130 HU)[25] as expected, since contrast-enhanced blood pool usually has an HU of 250-350.[47] Coronary artery and aortic valve calcium extraction from contrast-enhanced CT similarly uses higher derived thresholds (300-850 HU) for comparison with non-contrast scoring.[35, 48-50] We empirically evaluated non-contrast attenuation thresholds to identify the calcium voxel definition that best corresponded to our arterial-phase CTA volume measurements. In addition, our study shows good agreement with Pearson r>0.90 across a wide range of non-contrast ground truth thresholds.

This approach has several important application limitations: patients who have had stents placed or who have intraarterial devices cannot be accurately assessed, since the model cannot distinguish between stents and calcium. Calcium predictions may also not be accurate in patients with aneurysmal vessels with significant mural thrombus due to apparent reliance on contrast pool to define vessel bounds; we therefore recommend caution and manual review in these cases. Interestingly, the pipeline was able to handle short-segment arterial occlusions and accurately detect calcium within these regions, likely due to the nnU-Net learning an underlying representation of vessel geometry such that even missing contrast within the arterial lumen can be overcome (Supplemental Figure S1B). Finally, there are logistical barriers to the dissemination of this technique – those wishing to implement our techniques will need some computational resources and setup/approval within their healthcare system to use this tool.

Although we have demonstrated scalability between non-contrast ground truth and calcium volume predicted from contrast-enhanced scans and correlation between contrast and non-contrast calcium quartiles, there is imperfect congruence between these two measures. Contrast-enhanced calcium scores should not be directly leveraged in the same way as non-contrast scores until clinical interpretation thresholds are rigorously established. We plan to perform clinical validation of CTA-extracted aortoiliac calcium scores in our future work.

## Conclusion

Aortoiliac calcium burden can be accurately quantified from CTA using deep learning strategies and corresponds with calcium voxels ≥400 HU on non-contrast imaging. Automatic aortoiliac calcium segmentation may facilitate opportunistic detection of PAD and help quantify cardiovascular risk. The ability to extract calcium volume from arterial-phase, contrast-enhanced CT is the first step towards extending automatic calcium quantification to lower extremity CT angiograms, which could enable improved perfusion quantification and outcome prediction in PAD patients for whom ABIs are inaccurate.

## Data Availability

All data produced in the present study are available upon reasonable request to the authors

## Data and code availability

The imaging data are not publicly available because of Stanford University restrictions. The authors plan to release the code as open source following publication of the full work, but it is not yet publicly available.

## Sources of Funding

B.L. receives funding through the Stanford Cardiovascular Institute Boring Trust.

A.T.F. receives funding through T32HL166155 from the National Institutes of Health (NIH), Stanford Cardiovascular Institute, Computational Medicine in the Heart: Integrated Training Program.

A.F. and P.B. receive funding through T32EB009035 from the National Institutes of Health, Stanford Cardiovascular Institute, Multi-Disciplinary Training Program in Cardiovascular Imaging at Stanford.

F.R. was funded by grants from the NIH National Heart, Lung, and Blood Institute (R01HL168188; R01HL167974; R01HL169345) and the Doris Duke Foundation (Grant #2022051).

A.C. receives research support from NIH grants R01 HL167974, R01 HL169345, R01 AR077604, R01 EB002524, R01 AR079431, P41 EB027060; Advanced Research Projects Agency for Health (ARPA-H) contracts AY2AX000045 and 1AYSAX0000024-01; and NIH contracts 75N92020C00008 and 75N92020C00021. This work was also supported by Stanford seed grants from the Stanford Cardiovascular Institute, Stanford Center for Digital Health, Stanford Center for Artificial Intelligence in Medicine and Imaging, Stanford Institute for Human-Centered Artificial Intelligence, and Stanford Precision Health and Integrated Diagnostics.

S.A. is supported by grants from the American Heart Association (25TPA1479692) and the National Institutes of Health/National Heart, Lung, and Blood Institute (UG3HL181184, R01HL166305) and the Veterans Affairs Office of Research and Development (I01HX003215).

## Disclosures

F.R. reports consulting fees from Novartis, Esperion Therapeutics, Edwards, Arrowhead Pharmaceuticals, HeartFlow, iRhythm, Amgen, and Cleerly Health outside the submitted work.

Unrelated to this work, A.C. receives research support from GE Healthcare, Siemens, Philips, Microsoft, Amazon, Google, NVIDIA, Stability. A.C. has provided consulting services to Patient Square Capital, Chondrometrics GmbH, and Elucid Bioimaging; is co-founder of Cognita Imaging; receives consulting/advisory fees from Cognita Imaging; and has equity interest in Cognita Imaging, Subtle Medical, LVIS Corp, Brain Key, and Radiology Partners.

S.A. is a consultant for Gore Medical and Novartis Pharmaceutical outside the submitted work.

AI use statement: During the preparation of this work the authors used OpenAI ChatGPT-6.1 Sol to edit for syntax and readability. After using this tool, the authors reviewed and edited the content as needed and take full responsibility for the content of the publication.

## Notes

### Competing Interest Statement

The authors have declared no competing interest.

### Author Declarations

Institutional Review Board of Stanford University waived ethical ethical approval for this work (IRB 66789), due to entirely retrospective chart review nature of this research.

