## Supplementary for "Automated aortoiliac calcification segmentation from arterial-phase computed tomography scans: development and validation"

### Supplementary Section

#### *Generation of Training Data for ART nnU-Net*

SAM-2 was used to generate data for training ART nnU-Net. To adapt SAM-2 for volumetric analysis, we processed sequential axial CT slices as video frames. We initialized the segmentation mask at the L1 level using the TotalSegmentator model,[32] which served as a seed for the SAM-2 memory bank (Figure 3). Subsequent inference of the vessel segment was guided with logical rules attuned to the properties of the aortoiliac vasculature. To ensure exhaustive segmentation of the arterial tree, we employed a queue-based exploration strategy that initiated new tracking streams whenever a bifurcation was detected. Bifurcations were identified under two possible conditions: (1) a topological split resulting in disjoint mask components separated by more than 10 pixels, or (2) a geometric shift characterized by a >30% sudden reduction in cross-sectional area paired with a mask aspect ratio >1.5. To prevent the model from tracking visual artifacts, we terminated inference if the predicted mask vanished for 10 consecutive frames or exhibited implausible expansions defined as a >100% size increase relative to the rolling 10-frame average. Upon failure, the system utilized a backtracking mechanism, reverting 10 frames to reset the memory bank and restarting inference to circumvent anatomic abnormalities. After all inference streams converged, segment candidates were assembled into a directed graph. As a post-processing step, we employed a depth-first search (DFS) algorithm to reconstruct the final vessel volume by identifying the longest continuous path (bilateral) and thereby pruning superficial branches and visual tracking artifacts. The final pruned vessel volume was manually reviewed for inclusion in fully supervised training of the ART nnU-Net.

#### *Generation of Training Data for CA nnU-Net*

Comparison of bone and contrast-enhanced blood pool intensity was used to create arterial calcium training masks. We modeled the radiographic intensity distribution of calcium using the lumbar spine and the distribution of the luminal blood pool using the aortic core, defined as points excluding the outer 30% of the vessel's Euclidean distance transform profile. For every axial slice within the arterial volume segmented by ART nnU-Net, we initialized calcium segmentation predictions using a maximum likelihood estimation (MLE) approach. Voxels were classified as calcium if their intensity satisfied a log-likelihood ratio favoring the bone distribution over the blood pool distribution, adjusted by scaling the standard deviation of the bone distribution by a relaxation factor of 2.5. We refined our initial calcium mask predictions with an expansion into neighboring voxel regions using watershed segmentation.[51] We defined an "uncertainty zone" by dilating the initial calcium segmentation mask (3x3 voxel kernel, 2 iterations) and including voxels satisfying a looser MLE threshold (relaxation factor of 3.5). Within this region, we applied the watershed algorithm on the Sobel gradient magnitude[52] of the CT slice, using the high-confidence MLE predictions as foreground markers and the defined uncertainty zone as the unlabeled basin to be segmented. The resulting masks were then manually reviewed for inclusion in fully supervised training of the CA nnU-Net.

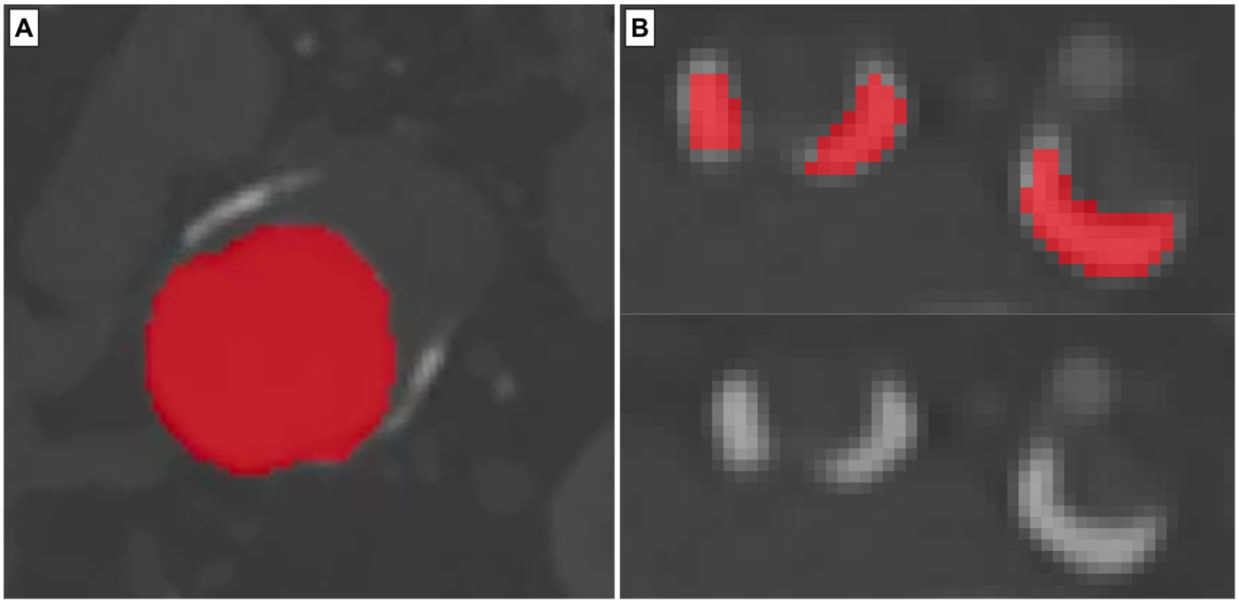


Figure S1: (A) Vessel segmentation (red) did not accurately extend to the aortic wall when significant mural thrombus was present, and arterial wall calcium was subsequently not captured for this area. (B) Calcium segmentation was accurately performed within bilaterally occluded common iliac artery segments (top with calcium segmentation overlay in red, bottom without segmentation overlay).

| | **ART nnU-Net (n = 38)** | **CA nnU-Net (n = 34)** | **Validation (n = 106)** |
|---|---|---|---|
| Female | 14 (37%) | 11 (32%) | 67 (63%) |
| Mean age (SD), years | 65.5 (14.3) | 68.8 (9.6) | 71.2 (17.6) |
| kVp (%) | | | |
| 70 - 90 | 7 (18%) | 5 (15%) | 12 (11%) |
| 100 - 110 | 11 (29%) | 9 (26%) | 30 (28%) |
| 120 - 140 | 20 (53%) | 20 (59%) | 64 (60%) |
| Slice thickness (%) | | | |
| 0.5-0.625 mm | 6 (16%) | 8 (24%) | 33 (31%) |
| 0.75-1 mm | 24 (63%) | 16 (47%) | 45 (42%) |
| 1.25-2.5 mm | 8 (21%) | 10 (29%) | 28 (26%) |

Table S1: Basic demographics and scan characteristics for all three cohorts used in the study: one training cohort each for arterial (ART) and calcium (CA) segmentation nnU-Net models, and one validation cohort for assessment of full pipeline outputs. Age is reported as mean (standard deviation [SD]) in years; other values are counts (percentages). kVp, peak kilovoltage. Percentages may not sum to 100% because of rounding.

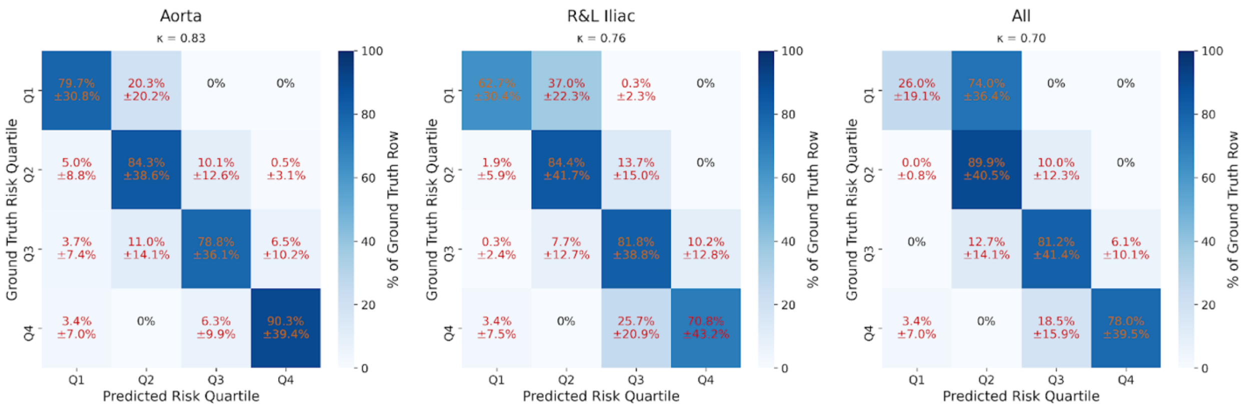


Figure S2: Linearly weighted kappa was calculated for the risk reclassification analysis. We found aortic κ = 0.83, bilateral iliac κ = 0.76, and all regions κ = 0.70.
